# Association between social determinants of health and cardiovascular and cancer mortality in cancer survivors: a nationally representative cohort study

**DOI:** 10.1101/2024.05.08.24307071

**Authors:** Jeffrey Shi Kai Chan, Danish Iltaf Satti, Yat Long Anson Ching, Quinncy Lee, Edward Christopher Dee, Kenrick Ng, Oscar Hou-In Chou, Tong Liu, Gary Tse, Agnes Lai

**Affiliations:** Cardio-Oncology Research Unit, Cardiovascular Analytics Group, PowerHealth Research Institute, Hong Kong, China; Division of Cardiology, Johns Hopkins University School of Medicine, Baltimore, MD, USA; Department of Radiation Oncology, Memorial Sloan Kettering Cancer Center, New York, NY, USA; Department of Medical Oncology, Barts Cancer Centre, London, UK; Tianjin Key Laboratory of Ionic-Molecular Function of Cardiovascular Disease, Department of Cardiology, Tianjin Institute of Cardiology, Second Hospital of Tianjin Medical University, Tianjin 300211, China; School of Nursing and Health Studies, Hong Kong Metropolitan University, Hong Kong, China; Kent and Medway Medical School, Canterbury, Kent, CT2 7NT, UK

**Keywords:** disparities, outcomes, epidemiology, cancer survivorship

## Abstract

Although it is known that social deprivation is associated with higher risks of cardiovascular and cancer mortality in the general population, these associations may not be directly applicable to cancer survivors. Using data from the National Health Interview Survey (NHIS) 2013-2017 with follow-up till the end of 2019, we examined the associations between social determinants of health (SDOH) and the risk of all-cause, cardiovascular, and cancer mortality in cancer survivors, with separate modelling in individuals without cancer for comparison. Two Cox regression models were fitted for each outcome, the first being adjusted for demographics and the second being additionally adjusted for comorbidities and risk factors. Altogether, 37,882 individuals were analysed (representing a weighted population of 57,696,771), including 4179 cancer survivors and 33,703 individuals without cancer. Amongst cancer survivors, worse SDOH was associated with higher all-cause, cardiovascular, and cancer mortality when adjusted for demographics, which were attenuated but remained significant when further adjusted for comorbidities and risk factors. Amongst individuals without cancer, worse SDOH was associated with higher all-cause and cardiovascular mortality only when adjusted for demographics, but not when further adjusted for comorbidities and risk factors. These suggested that in cancer survivors, associations between SDOH and cardiovascular/cancer mortality may be driven by factors beyond comorbidities and risk factors, contrasting individuals without cancer in whom similar associations were probably driven mostly by comorbidities and risk factors.

---

Social determinants of health (SDOH) are increasingly recognized to substantially influence cardiovascular health in both the general population and cancer survivors.^1,2^ A recent nationwide cross-sectional study of cancer survivors found strong associations between SDOH and cardiovascular risk factors.^3^ Despite known associations between SDOH and cardiovascular/cancer mortality in the general population,^4,5^ these associations have rarely been studied at the individual level amongst cancer survivors – given their cancer history and elevated cardiovascular risk,^6^ relevant associations observed in the general population may not be applicable to them. Additionally, few studies have explored SDOH comprehensively using composite metrics. Therefore, we explored associations between SDOH and cardiovascular and cancer mortality in cancer survivors using a published composite SDOH score. Individuals without cancer were also studied to explore whether these associations varied with cancer survivorship.

Methods are detailed in the **Supplementary Methods**. We used data from the National Health Interview Survey (NHIS), an annual survey representative of the United States’ non-institutionalized population linked to the National Death Index (NDI). Details of NHIS and data access were described elsewhere.^3,7,8^ All data underlying this study are publicly available, rendering it exempt from ethics review.

Participants in NHIS 2013-2017 with mortality follow-up data were included. Those with missing SDOH/covariate data were excluded. All subjects were followed up from questionnaire administration to the end of 2019 or death, whichever earlier, as detailed elsewhere.^9^ The outcomes of interest were all-cause mortality, cardiovascular mortality, and cancer mortality, ascertained through the NDI.

Cancer survivorship was self-reported^3,7^; per convention, individuals with only non-melanotic skin cancer were not considered cancer survivors.^3^ SDOH was quantified using a published, self-reported 38-point score, with higher scores indicating worse deprivation.^3^ Covariates were ascertained from self-reported data as previously detailed.^3,7^

Survey-specific statistics with sampling weights were used to produce nationally representative estimates. Due to right-skewing, the composite SDOH score was analysed as standardized continuous variables after log-transformation (i.e. ln[SDOH+1]; abbreviated as ‘SDOH’ hereafter). As non-cardiovascular-non-cancer mortality (‘other-cause mortality’) constituted a competing event for cardiovascular and cancer mortality, a cause-specific approach was adopted, modelling associations between SDOH and the risks of each outcome and other-cause mortality using Cox regression. Associations in cancer survivors and individuals without cancer were modelled separately. Two models were pre-specified for each outcome based on clinical knowledge: model 1 was adjusted for demographics, while model 2 was adjusted for demographics, comorbidities, and risk factors.

A total of 37,882 individuals were analysed (**Supplementary Figure 1**), representing a population of 57,696,771 persons after applying sampling weights. These included 4179 cancer survivors (weighted N=5,762,493) and 33,703 individuals without cancer (weighted N=51,934,278). Their characteristics were summarized in **Supplementary Table 1**.

The analytic results were summarized in **Figure 1** and **Supplementary Table 2**. Amongst cancer survivors, worse SDOH was associated with higher all-cause, cardiovascular, and cancer mortality when adjusted for demographics, which were attenuated but remained significant when further adjusted for comorbidities and risk factors. No significant associations were found between SDOH and other-cause mortality.

**Figure 1.**
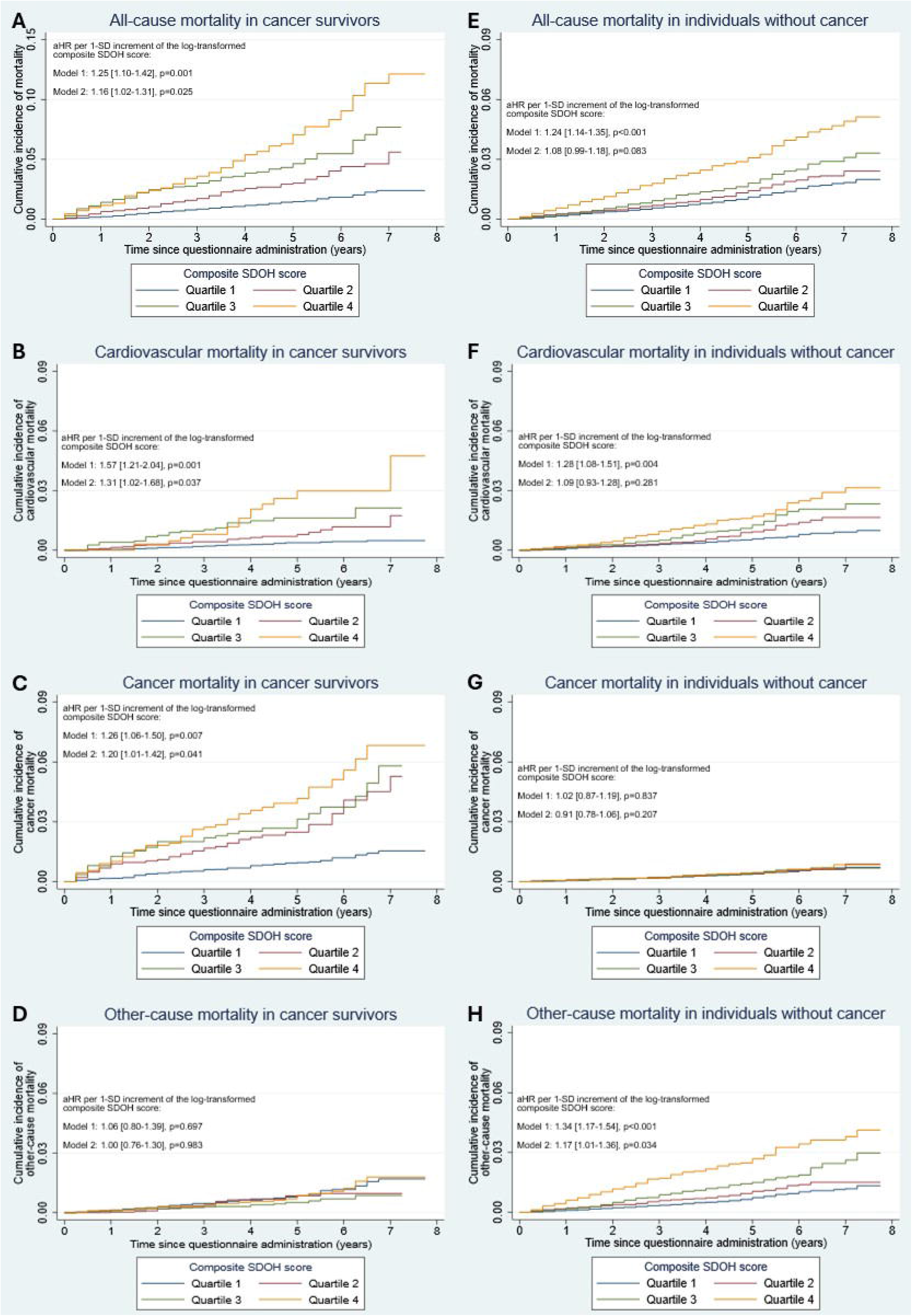
Kaplan-Meier cumulative incidence curves of all-cause (**A, E**), cardiovascular (**B, F**), cancer (**C, G**), and other-cause (**D, H**) mortality in cancer survivors (**A-D**, respectively) and individuals without cancer (**E-H**, respectively). All summary statistics presented were adjusted hazard ratios (aHR) with 95% confidence intervals. Model 1 was adjusted for age, race, and sex. Model 2 was adjusted for age, race, sex, hypertension, diabetes mellitus, dyslipidaemia, active smoking, weekly number of alcoholic drinks, cardiac condition(s), chronic obstructive pulmonary disease or emphysema, liver disease, stroke, obesity, and weekly exercise duration.

Meanwhile, amongst individuals without cancer, worse SDOH was associated with higher all-cause and cardiovascular mortality only when adjusted for demographics, but not when further adjusted for comorbidities and risk factors. Worse SDOH was associated with other-cause mortality in both multivariable models, whilst no associations between SDOH and cancer mortality were found.

This was one of the first studies specifically investigating associations between SDOH and cancer/cardiovascular mortality in cancer survivors. Our findings suggested that in cancer survivors, associations between SDOH and cardiovascular/cancer mortality may be driven by factors beyond comorbidities and risk factors which *per se* are associated with SDOH,^3^ whilst much of the SDOH-cardiovascular mortality association in subjects without cancer was driven by comorbidities and risk factors. This highlighted the importance of addressing SDOH to improve cardiovascular/cancer outcomes in cancer survivors. Speculatively, disparity in multidisciplinary cardio-oncology care access may have driven our findings for cardiovascular mortality in cancer survivors, warranting relevant research. Whether the observed SDOH-mortality associations differ by race/ethnicity also warrants further research.^1,10^

Notwithstanding this study’s nationally representative nature, it was limited by NHIS’ self-reported nature, and the lack of individual data adjudication potentially predisposing to mortality data miscoding. Low event rates precluded subgroup/exploratory analyses, and the observational nature precluded causal inferences. Overall, our findings are hypothesis-generating, necessitating further confirmatory/mechanistic studies.

In conclusion, amongst cancer survivors, worse SDOH was associated with higher cardiovascular and cancer mortality, independent of demographics, comorbidities, and risk factors. Amongst individuals without cancer, SDOH was associated with cardiovascular mortality independent of demographics but not comorbidities and risk factors, with no association between SDOH and cancer mortality found. Further confirmatory/mechanistic studies are required.

## Data Availability

All data underlying this study are publicly available and accessible online at https://nhis.ipums.org/nhis/.

