## Supplementary for "Association between social determinants of health and cardiovascular and cancer mortality in cancer survivors: a nationally representative cohort study"

**SUPPLEMENTARY MATERIAL**

**Supplementary methods**

Data from the National Health Interview Survey (NHIS) were used. The NHIS, an annual survey of the United States' non-institutionalized population linked to the National Death Index (NDI), uses multistage probability sampling to generate nationally representative estimates. Details of NHIS and data access were described elsewhere.[1–3] All data underlying this study are publicly available.[3] Therefore, this study is exempt from ethics review.

Participants in NHIS 2013-2017 with mortality follow-up data were included – only these years/iterations contained variables necessary for quantifying SDOH. Those with missing SDOH or covariate data were excluded. All subjects were followed up from questionnaire administration to the end of 2019 or death, whichever occurred earlier, as detailed elsewhere.[4] The outcomes of interest were all-cause mortality, cardiovascular mortality, and cancer mortality, ascertained through the NDI using death certificate information.

Cancer survivorship was self-reported[1,2]; per convention, individuals with only non-melanotic skin cancer were not considered cancer survivors.[1] SDOH was quantified using a published, self-reported 38-point score, with higher scores indicating worse deprivation.[1] Covariates, including demographics (age, race, and sex) and comorbidities / risk factors (hypertension, diabetes mellitus, hypercholesterolemia, active smoking, obesity, chronic obstructive pulmonary disease or emphysema, stroke, weekly moderate/vigorous exercise duration, weekly number of alcoholic drinks, and cardiac and liver conditions), were ascertained from self-reported data as previously detailed.[1,2]

Survey-specific statistics with sampling weights were used via Stata's *svy* set of commands to produce nationally representative estimates. Due to the survey nature of the data, continuous variables were summarized as means and 95% confidence intervals (CIs), while categorical variable were summarized as proportions and 95% CIs. As individuals with missing data were excluded, there were no missing data in this study.

Due to right-skewing, the composite SDOH score was analysed as standardized continuous variables after log-transformation (i.e. ln[SDOH+1]; abbreviated as 'SDOH' hereafter). As non-cardiovascular-non-cancer mortality ('other-cause mortality') constituted a competing event for cardiovascular and cancer mortality, a cause-specific approach was adopted, modelling associations between SDOH and risks of each outcome and other-cause mortality using Cox regression. Log-log plots showed no violation of the proportional hazard assumption. Kaplan-Meier curves were used to visualize the cumulative incidence of each outcome and other-cause mortality, with grouping by quartiles of the SDOH score – SDOH was quantitatively analysed as a continuous variable instead of quartiles as the latter reduced the sample size, event rate, and thereby the power of Cox regression substantially, which rendered quartile-based analysis infeasible given the already-low event rates.

Associations in cancer survivors and individuals without cancer were modelled separately. Two models were pre-specified for each outcome based on clinical knowledge: model 1 was adjusted for demographics, while model 2 was adjusted for demographics, comorbidities, and risk factors.

Two-sided $p<0.05$ were considered statistically significant. All analyses were performed using Stata v16.1 (StataCorp LLC, College Station, Texas, USA).

**Supplementary Table 1.** Characteristics of analysed individuals. Continuous variables were summarized as means and 95% confidence intervals (CIs). Categorical variables were summarized as proportions and 95% CIs.

| | Individuals without cancer | Cancer survivors |
|---|---|---|
| Sample size | 33,703 | 4179 |
| Weighted sample size | 51,934,278 | 5,762,493 |
| ***Demographic and comorbid characteristics****, mean [95% CI] or proportion (%) [95% CI (%)]* | | |
| Age, years old | | |
| 18-25 | 9.4 [8.8-9.9] | 0.8 [0.4-1.3] |
| 26-35 | 16.6 [16.1-17.2] | 2.9 [2.3-3.6] |
| 36-45 | 17.4 [16.9-17.9] | 6.2 [5.3-7.3] |
| 46-55 | 20.5 [19.9-21.1] | 14.7 [13.2-16.3] |
| 56-65 | 19.9 [19.3-20.5] | 24.1 [22.4-25.9] |
| 66-75 | 11.5 [11.1-12.0] | 30.6 [28.8-32.3] |
| ≥76 | 4.7 [4.4-5.0] | 20.8 [19.3-22.3] |
| Male | 48.8 [48.1-49.5] | 49.6 [47.6-51.6] |
| Race | | |
| White | 85.4 [84.7-86.1] | 92.2 [91.1-93.2] |
| Black / African American | 9.2 [8.7-9.7] | 5.7 [4.9-6.7] |
| American Indian / Alaskan native | 0.8 [0.7-1.0] | 0.5 [0.3-0.7] |
| Chinese | 0.9 [0.7-1.0] | 0.2 [0.1-0.7] |
| Filippino | 1.1 [0.9-1.3] | 0.4 [0.3-0.8] |
| Asian Indian | 0.9 [0.7-1.1] | 0.1 [0.0-0.2] |
| Other Asians | 1.3 [1.1-1.5] | 0.5 [0.3-0.8] |
| Other / multiple races | 0.4 [0.4-0.5] | 0.3 [0.2-0.6] |
| Hypertension | 38.4 [37.7-39.1] | 53.9 [51.8-56.0] |
| Diabetes mellitus | 12.8 [12.3-13.2] | 20.4 [18.8-22.1] |
| Hypercholesterolemia | 36.1 [35.4-36.8] | 51.6 [49.6-53.7] |
| Active smoking | 42.4 [41.6-43.2] | 55.0 [52.9-57.1] |
| Obesity | 32.9 [32.2-33.6] | 31.3 [29.4-33.1] |

| | Individuals without cancer | Cancer survivors |
|---|---|---|
| Cardiac condition | 12.5 [12.0-12.9] | 25.8 [24.1-27.5] |
| Liver condition | 2.2 [2.0-2.4] | 4.4 [3.7-5.3] |
| Chronic obstructive pulmonary disease or emphysema | 3.3 [3.0-3.5] | 8.5 [7.5-9.7] |
| Stroke | 2.3 [2.1-2.5] | 5.2 [4.4-6.2] |
| Weekly moderate/vigorous exercise duration, minutes | 262 [256-268] | 238 [220-256] |
| Weekly number of alcoholic drink(s) | 4.7 [4.4-4.9] | 4.8 [4.4-5.2] |
| Composite social determinants of health score | 6.0 [5.9-6.1] | 5.3 [5.2-5.5] |
| Log-transformed composite social determinants of health score | 1.75 [1.74-1.76] | 1.62 [1.59-1.65] |
| ***Domains of the composite social determinants of health score****, proportion (%) [95% CI (%)]* | | |
| *Economic stability* | | |
| Never / previously employed | 3.0 [2.7-3.2] | 1.6 [1.2-2.2] |
| No paid sick leave | 37.1 [36.4-37.9] | 36.9 [34.9-38.9] |
| Low family income | 18.1 [17.5-18.8] | 16.4 [15.1-17.8] |
| Difficulty paying medical bills | 12.9 [12.3-13.4] | 11.9 [10.7-13.3] |
| Unable to pay medical bills | 6.2 [5.8-6.5] | 5.3 [4.5-6.2] |
| Cost-related medication non-adherence | 8.7 [8.3-9.1] | 8.9 [7.9-10.1] |
| Foregone / delayed medical care due to cost | 9.2 [8.8-9.6] | 7.4 [6.5-8.5] |
| Worried about money for retirement | 47.5 [46.7-48.2] | 37.9 [36.0-39.9] |
| Worried about medical costs of illness / accident | 41.8 [41.0-42.6] | 33.8 [32.0-35.7] |
| Worried about maintaining standard of living | 37.4 [36.7-38.1] | 33.5 [31.7-35.4] |
| Worried about medical costs of normal healthcare | 25.3 [24.6-26.0] | 21.8 [20.1-23.5] |
| Worried about paying monthly bills | 24.3 [23.6-25.0] | 21.8 [20.2-23.5] |
| Worried about paying rent / mortgage / housing costs | 18.6 [18.0-19.2] | 15.9 [14.5-17.5] |
| *Neighborhood, physical environment, and social cohesion* | | |
| Housing was rental / from other arrangement | 27.2 [26.4-28.0] | 16.7 [15.4-18.2] |
| People in neighborhood did not help each other | 14.4 [13.9-14.9] | 11.7 [10.5-13.0] |
| There were not people that can be counted on in neighborhood | 14.6 [14.1-15.2] | 10.9 [9.7-12.1] |
| People neighborhood could not be trusted | 33.6 [32.8-34.3] | 31.0 [29.3-32.8] |
| Neighborhood was not close-knit | 12.9 [12.3-13.4] | 9.5 [8.3-10.7] |

| | Individuals without cancer | Cancer survivors |
|---|---|---|
| *Psychological distress* | 3.3 [3.0-3.6] | 3.3 [2.7-4.1] |
| *Food insecurity* | 6.5 [6.1-6.8] | 5.9 [5.0-6.8] |
| *Education* | | |
| Could not speak English language well / at all | 1.7 [1.5-1.9] | 1.3 [0.9-1.8] |
| Did not look up health information on internet in the past 12 months | 64.7 [63.9-65.4] | 59.3 [57.3-61.3] |
| Did not fill a prescription on the internet in the past 12 months | 16.4 [15.8-17.0] | 17.3 [15.8-18.9] |
| Did not schedule medical appointment on the internet in the past 12 months | 17.2 [16.5-18.0] | 15.3 [13.8-16.9] |
| Did not communicate with healthcare provider by email in the past 12 months | 19.0 [18.3-19.8] | 21.0 [19.4-22.7] |
| Did not use chat groups to learn about health topics in the past 12 months | 4.7 [4.4-5.0] | 5.4 [4.5-6.5] |
| Less than high school education | 25.9 [25.2-26.7] | 27.5 [25.8-29.2] |
| *Healthcare system* | | |
| Uninsured | 5.3 [5.0-5.7] | 2.5 [1.9-3.1] |
| No usual source of care | 6.2 [5.8-6.5] | 2.9 [2.4-3.7] |
| Trouble finding a doctor / healthcare provider | 3.0 [2.8-3.3] | 3.7 [3.0-4.6] |
| Not accepted by doctor's office as new patient | 2.8 [2.6-3.1] | 3.5 [2.8-4.3] |
| Insurance not accepted by doctor's office | 3.8 [3.6-4.2] | 4.3 [3.6-5.2] |
| Delayed medical care due to not being able to get through on the phone | 3.1 [2.8-3.4] | 3.2 [2.6-3.9] |
| Delayed medical care due to not being able to get an appointment soon enough | 7.9 [7.5-8.4] | 8.5 [7.4-9.7] |
| Delayed medical care due to waiting too long at the doctor's office | 4.2 [3.9-4.5] | 5.4 [4.6-6.4] |
| Delayed medical care due to the doctor's office not being open when there was time to visit | 3.7 [3.4-4.0] | 3.0 [2.4-3.8] |
| Delayed medical care due to a lack of transportation | 1.5 [1.3-1.6] | 2.1 [1.6-2.7] |
| Dissatisfied with the quality of care / no healthcare in the past year | 6.6 [6.2-6.9] | 5.1 [4.3-6.1] |

CI, confidence interval.

**Supplementary Table 2**. Summary of follow-up duration, outcomes, competing event, and analytic results.

| | Individuals without cancer | Cancer survivors |
|---|---|---|
| *Follow-up duration, outcomes, and competing event, mean [95% CI] or proportion (%) [95% CI (%)]* | | |
| Follow-up duration, years | 4.8 [4.8-4.8] | 4.6 [4.6-4.7] |
| All-cause mortality | 2.4 [2.2-2.6] | 9.9 [8.8-11.0] |
| Cardiovascular mortality | 0.7 [0.6-0.8] | 2.2 [1.8-2.8] |
| Cancer mortality | 0.5 [0.4-0.6] | 4.6 [3.9-5.5] |
| Other-cause mortality | 1.1 [1.0-1.2] | 3.0 [2.4-3.7] |
| *Association between the standardized log-transformed composite SDOH score and risk of outcomes, adjusted hazard ratio [95% CI]** | | |
| All-cause mortality | Model 1**: 1.24 [1.14-1.35], p<0.001<br>Model 2***: 1.08 [0.99-1.18], p=0.083 | Model 1**: 1.25 [1.10-1.42], p=0.001<br>Model 2***: 1.16 [1.02-1.31], p=0.025 |
| Cardiovascular mortality | Model 1**: 1.28 [1.08-1.51], p=0.004<br>Model 2***: 1.09 [0.93-1.28], p=0.281 | Model 1**: 1.57 [1.21-2.04], p=0.001<br>Model 2***: 1.31 [1.02-1.68], p=0.037 |
| Cancer mortality | Model 1**: 1.02 [0.87-1.19], p=0.837<br>Model 2***: 0.91 [0.78-1.06], p=0.207 | Model 1**: 1.26 [1.06-1.50], p=0.007<br>Model 2***: 1.20 [1.01-1.42], p=0.041 |
| Other-cause mortality | Model 1**: 1.34 [1.17-1.54], p<0.001<br>Model 2***: 1.17 [1.01-1.36], p=0.034 | Model 1**: 1.06 [0.80-1.39], p=0.697<br>Model 2***: 1.00 [0.76-1.30], p=0.983 |

CI, confidence interval.

* Adjusted hazard ratios represented effects per standard deviation increase in the log-transformed composite social determinants of health score.

** Model 1: adjusted for age, race, and sex.

*** Model 2: adjusted for age, race, sex, hypertension, diabetes mellitus, dyslipidaemia, active smoking, weekly number of alcoholic drinks, cardiac condition(s), chronic obstructive pulmonary disease or emphysema, liver disease, stroke, obesity, and weekly exercise duration.

**Supplementary Figure 1.** Participant flow diagram. Weighted N refers to the population represented by the respective sample cohort after applying sampling weights. NHIS, National Health Interview Survey. SDOH, social determinants of health.

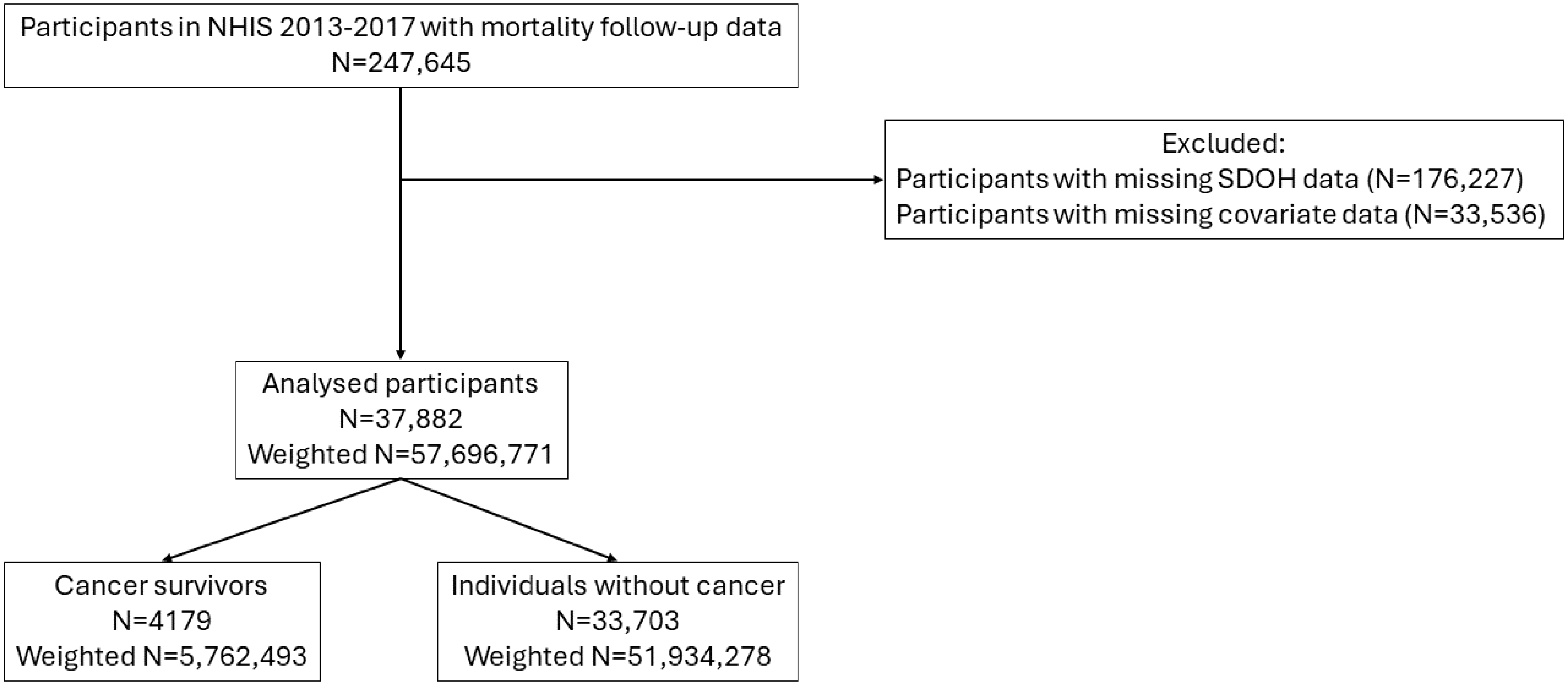
